# Reopening Universities without Testing During COVID-19: Evaluating a Possible Alternative Strategy in Low Risk Countries

**DOI:** 10.1101/2021.03.02.21250607

**Authors:** Jing Yang (Sunny) Xi, Wai Kin (Victor) Chan

**Affiliations:** Tsinghua-Berkeley Shenzhen Institute, Tsinghua University, Shenzhen, 518055, China

**Keywords:** Agent-based simulation, SEIR, COVID-19, reopen university, nonpharmaceutical intervention, low risk country

## Abstract

The safety of students worldwide remains a key issue during COVID-19. The reopening of universities in high risk countries during Fall 2020 resulted in numerous outbreaks. While regular screening and testing on campus can prevent the transmission of SARS-CoV-2, they are extremely challenging to implement due to various reasons such as cost and logistics. However, for low risk countries with minimal to no community spread, our study suggests that universities can fully reopen without testing, if students self-quarantine for 14 days on arrival and adopt proper nonpharmaceutical interventions (NPIs). This alternative strategy might save institutions millions of dollars. We adopt agent-based simulation to model virus transmission on campus and test the effectiveness of several NPIs when school reopens. Assuming one initially infected student, results indicate that transmission between roommates causes the most infections with visitors, ground floors, and elevators, being the next main contributors. Limiting density and/or population are not impactful at flattening the curve. However, adopting masks, minimizing movement, and increasing the frequency of cleaning can effectively minimize infection and prevent outbreak, allowing for classes and activities to resume as normal.

## 1 Introduction

As of February 2021, 28 countries still have yet to reopen schools, while others have partially opened or fully opened^1^. Prolonged school closure due to COVID-19 severely impacts students’ mental health, academic performance, social network, and family life^2^. However, reopening schools puts students, staff, and faculty at high risk^3^. Since universities and colleges in many countries reopened during Fall 2020, a number of them have experienced outbreaks on campus and become COVID-19 hot spots^4-9^. In the U.S. for instance, over a third of colleges and universities reopened fully in August^3^. By December, more than 397,000 cases and at least 90 deaths were documented across 1,900 schools^10^. These institutions of higher education are at extremely high risk of COVID-19 outbreak due to high density, high percentage of non-local students, and on-campus dormitories^3,11^.

In theory, regular screening and testing on university campus can facilitate reopening by detecting and preventing the transmission of SARS-CoV-2^12^. A number of studies have focused on their implementation in very high risk countries (as defined by the U.S. CDC^13-14^), such as in the U.S. and the U.K., where they are crucial for safe reopening, and provided related policy recommendations. Gillam et al. surveyed the feasibility and students’ acceptance of asymptomatic testing on a university campus^15^. Denny et al. discussed the implementation of a pooled surveillance testing program for asymptomatic SARS-CoV-2 infections in Duke university^12^. Ghaffarzadegan used equation-based model to investigate how testing, masking, enhanced communication, and remote work can help contain the spread of virus on campus^16^. Gressman and Peck developed stochastic agent-based model to determine the viability of in-person instruction when randomized testing, contact-tracing, and quarantining are systematically implemented^17^.

However, there are several limitations associated with regular testing, namely sensitivity, cost, and availability. Paltiel et al. estimated the testing cost for one semester of 80 days and 5,000 students using tests with 70%, 80%, and 90% sensitivity, respectively^18^. In the best-case scenario, weekly testing with 70% sensitivity would cost the school almost $700,000, whereas daily testing with 90% sensitivity would cost more than $20 million. Most institutions simply cannot financially afford and maintain regular testing of this magnitude. In addition, some health professionals pointed out that testing capacity can be severely limited at times, and results might take more than two weeks to arrive in countries facing large number of cases^19^. All of these issues deter the implementation of regular testing on campus.

Despite the theoretical promises tests bring in computer models, many universities that reopened in Fall 2020 lacked organized testing programs and policies; even for those that did, many only adopted symptom-based screening, which was not a robust prevention strategy^3^. For U.S. colleges with in-person classes and more than 5,000 students, only 25% conduct mass screening or random “surveillance” of students and only 6% regularly test all their students^12^. Past events have made evident that reopening universities in very high risk countries can lead to outbreaks on campus^20^, even if some form of testing policy is implemented. Therefore, we turn our attention to facilitating the safe reopening of universities in low risk countries, such as in Australia, China, and Greenland (as of May 2021 defined by the U.S. CDC^14^) where community spread is minimal or nonexistent.

The objective of this study is to model and assess an alternative strategy that do not utilize testing for university reopening in low risk countries. Instead, students will self-quarantine in their residences for 14 days before classes start. This approach is adopted by our school as well as others^12,21,22^ around the world and is shown to reduce test positivity rates among students^2^. In addition, students and campus administrators will adopt several nonpharmaceutical interventions (NPIs) to prevent the spread of SARS-CoV-2. If universities in low risk countries can safely reopen without testing due to low introduction risk^23^ of COVID-19, they can potentially save millions in testing costs. This prospect has not been explored in previous literatures as most of them focus on very high risk countries where testing and screening are required. To achieve our objective, we design an agent-based SEIR model of our campus and simulate the daily routines and health states of 2,952 residential students during this self-quarantine period. We empirically validate our model by recreating two previous outbreaks in university dormitories and comparing our simulation results against historic data. Assuming one initially infected student, we track the modes of virus transmission on campus and test six NPIs commonly used in schools^24^. The effectiveness and practicality of each NPI are discussed.

This study provides several contributions in an effort to prevent the spread of COVID-19 in universities. First, we propose an agent-based model to simulate virus transmission dynamics in a university dormitory setting to identify the main avenues of infection. Second, we implement six different NPIs at the individual level to assess their effectiveness. Third, we mathematically simulate and validate the viability of the 14 days quarantine strategy adopted by schools around the world. Finally, we suggest an optimal strategy for the reopening of universities in low risk countries that maximizes students’ safety, convenience of living, and access to education.

## 2 Methods

To simulate the transmission of SARS-CoV-2 between residential students on our campus, we propose an agent-based SEIR (Susceptible, Exposed, Infectious, Removed) model. At the micro level, each student is an agent capable of interaction and possess a health state of either “S”, “E”, “I”, or “R”, which will change when they get infected. At the macro level, the total numbers of students with “S”, “E”, “I”, and “R” are tallied, generating a stochastic compartmental model for the entire school.

### 2.1 Model Setup

On campus, there are four dormitory buildings, two 17-story buildings each with two elevators and two 26-story buildings each with three elevators. The ground floor (1^st^ floor) is uninhabited by students. Each floor above hosts 12 rooms, with a maximum occupancy of three students. In total, 2,952 student agents are organized into these buildings. Moreover, there are two canteen buildings with a total of 15 windows where students line up to purchase food.

Our model is constructed in NetLogo^25^ where all students arrive on day 1 from different cities across the country and undergo 14 days of quarantine in their residences on campus. During this time, students are restricted to their residences and canteens. We assume other people on campus, such as faculty and staff do not have enough interactions with students to play a role in the virus transmission during this period. In addition, there is no migration of any student in or out of campus, except when taken to hospital for treatment. Each simulation starts with one infected student and ends after eight weeks. Each time iteration, or tick, represents 30 minutes real-time.

### 2.2 Modes of Transmission

Every day during breakfast (7:00-9:00), lunch (11:00-13:00), and dinner (17:00-19:00), students go to the window of their choices to purchase food for takeout as dine-in is not permitted. Since university students are quite social, on each day, a percentage of all students will feel like visiting their friends. Each student planning to visit travels to the room of a randomly chosen student. We assume infections can only occur inside enclosed spaces in the canteens and dormitories and cannot occur when students walk outside between different buildings. With the current infrastructure setup, we identify six possible ways of infection, where *τ* represents transmissibility (table 1).

**Table 1.** Modes of virus transmission on campus.

|  | Mode |  | Transmissibility |  |
| --- | --- | --- | --- | --- |
|  | Name | Maintain social distancing? | Between students (direct) | Between student and environment (indirect) |
| 1. | Roommate | No | 100% (prolonged face-face) | N/A |
| 2. | Visitor | No | 100% (prolonged face-face) | N/A |
| 3. | Elevator | No | $\tau$ | $\tau$ |
| 4. | Hallway | Yes | N/A | $\tau$ |
| 5. | Ground Floor | Yes | N/A | $\tau$ |
| 6. | Canteen (no dine-in) | Yes | N/A | $\tau$ |

Dormitory rooms are small, poorly circulated, and almost never cleaned, a perfect environment for the virus to spread. Prolonged face-to-face contact with an infected individual in this environment would most likely result in healthy students being infected. Therefore, we use 100% chance to represent guaranteed viral transmission between students in the same room. Each time in a public area, an “I” student has a chance (*τ*) to infect their surrounding environment; an “S” student has the same chance of getting infected from this environment, if contaminated. We assume students would practice sufficient social distancing when walking in their buildings or waiting in lines at the canteens. As a result, no direct transmission can occur in public areas, except for in elevators, where it is impossible to maintain a safe distance. There are other possible modes for SARS-CoV-2 to transmit, such as through fecal aerosol or airborne transmission between flats in high-rise buildings^26-28^, however, they are not included in our simulation.

### 2.4 Estimating Transmissibility *τ*

At the micro level, the health state of each student is represented by either “S”, “E”, “I”, or “R” (figure 1). We assume no immunity, asymptomatic patient, or death. When an “S” student gets infected, they get assigned an incubation period, *i*, and duration of infection, *d*, randomly generated according to table 2. All epidemiological parameters in our model are compiled from previous literatures, except for *τ* and *c*, which are parameters specific for agent-based models, that is, their values depend on model setup.

**Table 2.** Epidemiological parameters of SARS-CoV-2 compiled from literature.

| Parameter | Value | Source |
| --- | --- | --- |
| Basic reproduction number, $R_0$ | 3.28 (1.5 to 6.49) | [29] |
| Incubation period, $i$ | lognormal (5.0) | [29] |
| Duration of infection, $d$ | 2.3 (0.8 to 3.0) | [30] |
| Latent period, $l$ | $i - d = 2.7$ | [30] |
| Infection rate, $\beta$ | $R_0/d = 3.28/2.3$ | [31] |
| Removal rate, $\gamma$ | $d^{-1} = 1/2.3$ | [32] |
| Onset rate, $\alpha$ | $l^{-1} = 1/2.7$ | [32] |
| Transmissibility, $\tau$ | 0.0082 | Model calibration |
| Average contact rate, $c$ | $R_0/d/\tau = 174$ | [31] |

**Figure 1.**
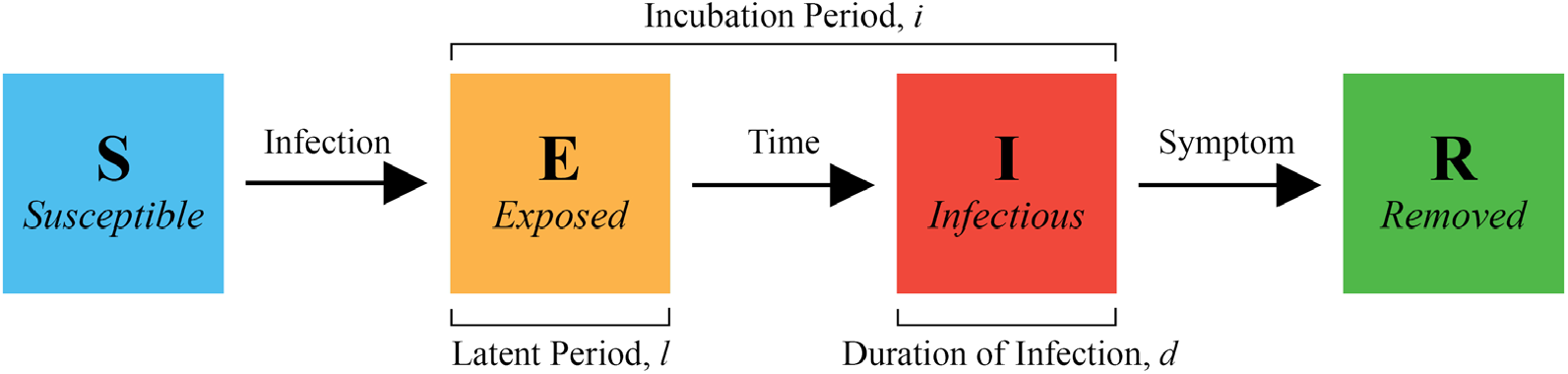
Conditions for changing health states. When an “S” student is infected, they change to “E”, and the model randomly generates an incubation period and duration of infection. With time, “E” transitions to “I” and “R” with symptom onset. “R” students are removed from campus for isolation or treatment and can no longer cause infections.

We estimate *τ* by plotting our stochastic model against the classic deterministic SEIR model^31^ using parameters listed in table 2 and subject to model initial conditions. The three key parameters for the SEIR model are *β, α*, and *γ*. A similar approached is adopted in^33^ for an SIR model. *τ* is calibrated to be 0.82% (figure 2) and the model is validated by comparing simulated outbreaks to previous outbreaks observed in universities (figure 3).

**Figure 2.**
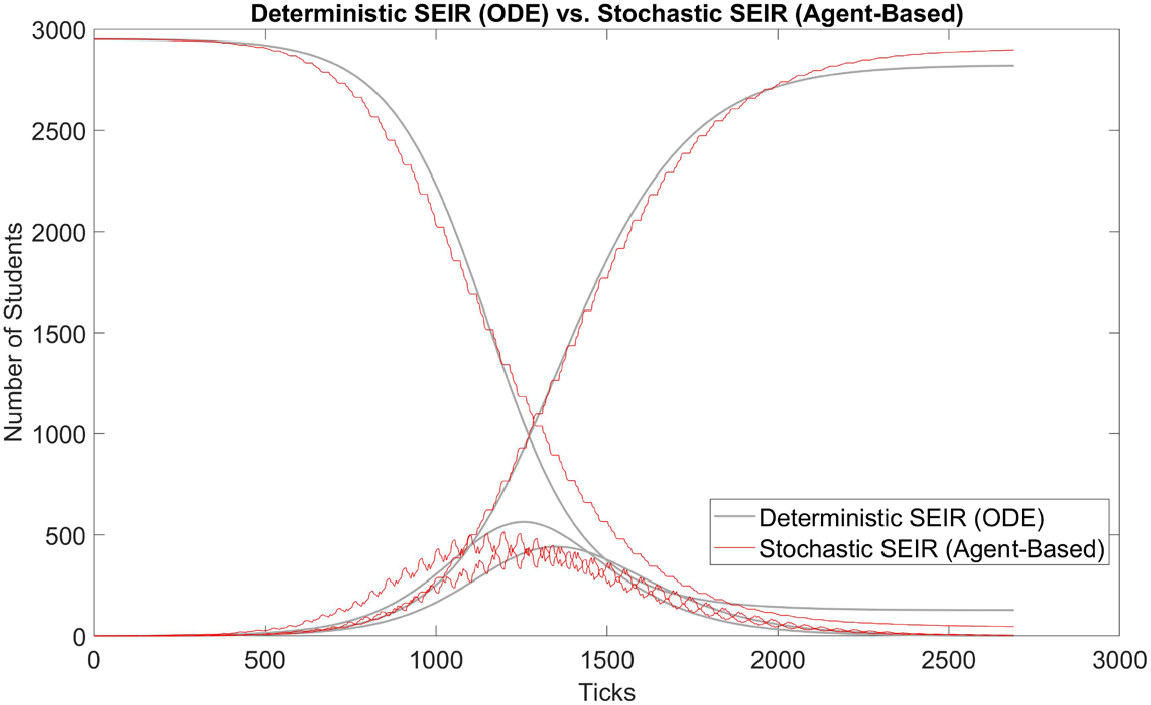
Model calibration: comparison between deterministic and stochastic SEIR (our model, 100 runs) with *τ* = 0.82%. The x-axis is in simulation ticks, where each tick represents 30 mins of real time. The shark teeth pattern of the stochastic curves reflects the daily routine of students. Each incline in E(t) represents students are awake and infecting others, increasing the number of “E” students; each decline represents students are asleep, and the number of “E” students decreases as some of them transition to the “I” state. Due to the two states of awake and asleep, E(t) and I(t) are 180 degrees out of phase.

**Figure 3.**
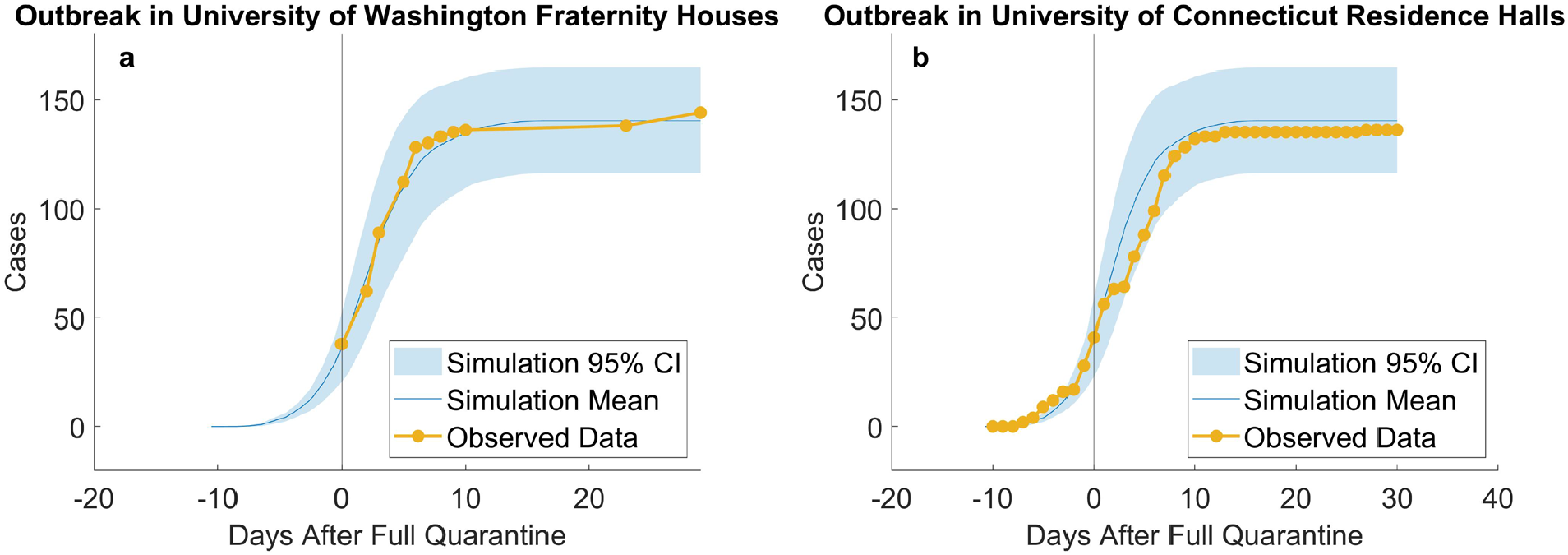
Empirical validation: simulating outbreaks in University of Washington fraternity houses (a, 10 runs) and University of Connecticut residence halls (b, 10 runs). According to reports from the University of Washington, “as of June 30^th^, at least 38 students living in 10 fraternity houses have tested positive”^34^. In total, there were about 1,000 residents in 25 fraternity houses and residents were asked to quarantine or self-isolate. We assumed that full quarantine started on that day, that is, no more uninfected residents could be infected, and used our model to simulate 1,000 students. Our results are plotted against observed data from June 30^th^ to Aug. 5^th^. A similar outbreak^35-36^ took place in University of Connecticut’s Storrs campus where five residence halls containing 544 students went into full quarantine starting Nov. 11^th^. On UConn’s COVID dashboard^37^, we compiled the daily number of new cases from 10 days before and 30 days after full quarantine and compared them to our simulated replication. Results suggest that our model provide close estimations to real outbreaks in university residences.

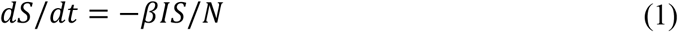

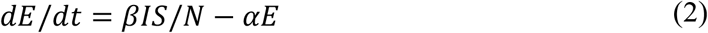

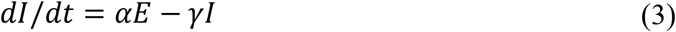

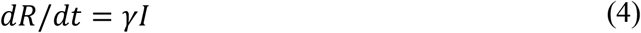

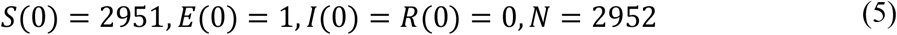

### 2.5 Experiments

The simulation starts with one initially infected student (*N*_*1*_ = 1), capable of infecting the entire campus if not controlled. Fortunately, we identify six possible NPI strategies^38^ that could slow down the speed of new infections and decrease the number of total infections (table 3). For each strategy, we repeat the simulation 100 times. The combination of red numbers in each row represents the baseline scenario, a typical daily routine without any active COVID-19 intervention measures.

**Table 3.** Six NPI strategies and their ranges of values (baseline values are in red).

| NPI Strategy | Variable affected | Range of Values |
| --- | --- | --- |
| 1. Adopt surgical masks: approx. 68% protection rate<sup>39</sup> | percent of students who use masks | [0% 25% 50% 75% 100%] |
| 2. Sanitize common areas more frequently | frequency of daily sanitizations | [0 1 2 3 4] |
| 3. Limit unnecessary movements | percent of students who visit friends | [0% 2.5% 5% 7.5% 10%] |
| 4. Limit max. number of students in an elevator | elevator allowance | [2 4 6 8] |
| 5. Limit number of students in each room | density | [1 2 3] |
| 6. Limit total number of students on campus | population | [50% 100% 200%] of 2952 |

## 3. Results

Based on simulation data, our end results could be categorized into runs with outbreak and runs without outbreak, as shown in figure 4. We define a new metric called outbreak probability to quantify the risk of uncontained outbreak on campus.

**Figure 4.**
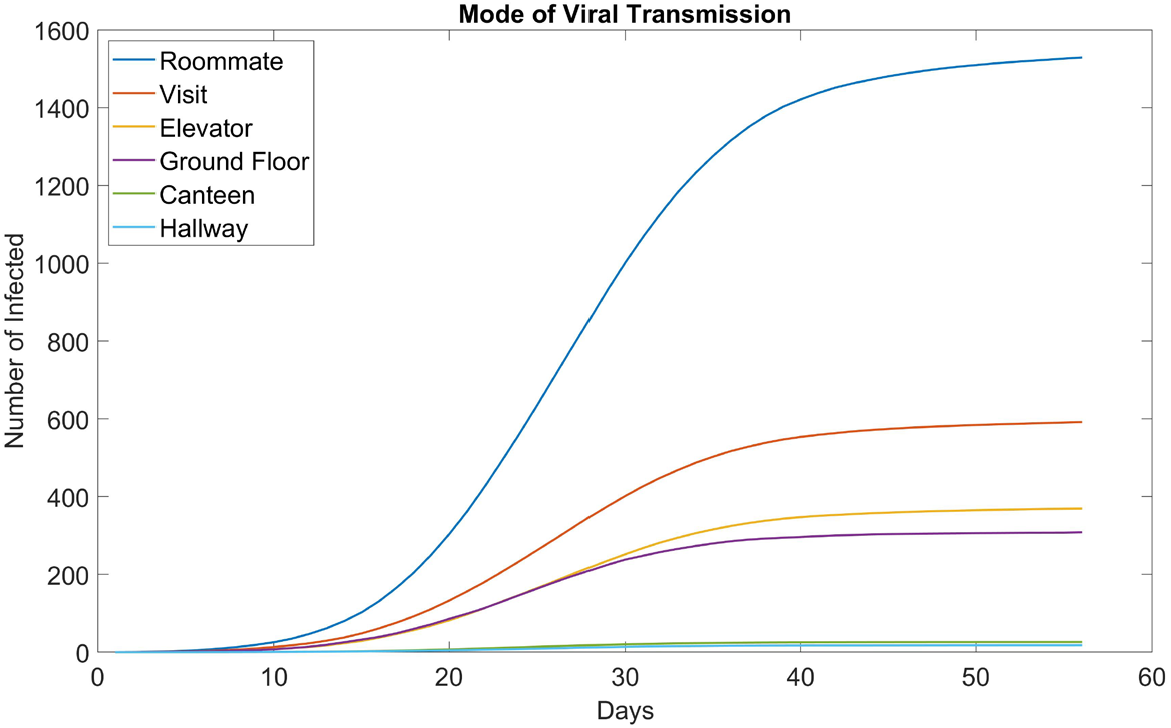
Histogram of total number of infected in the baseline scenario (100 runs). Due to the stochastic nature of our model, most end results are either located on the very left (no outbreak) or the very right (full outbreak). To be more exact, 49 out of 100 runs ended up with less than 30 infected students, or approximately 1% of the initial population *N*. In these runs, the infection in the campus is contained early on and do not end in an outbreak.

**DEFINITION 1**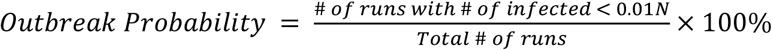

This bipolar phenomenon and its associated threshold are explored further and experimented on by Orbann et al^40^. From this point on, runs that do not result in outbreak are excluded from figures. Runs that do result in outbreak are averaged into one single timeseries and their frequencies are indicated.

### 3.1 Mode of Virus Transmission

Figure 5 shows how many students are infected in each location for the baseline scenario. Unsurprisingly, the largest contributor is roommate, accounting for over half the cases. This is troubling because if the school wants to fully resume functionality, it is impossible for students to not have roommates.

**Figure 5.**
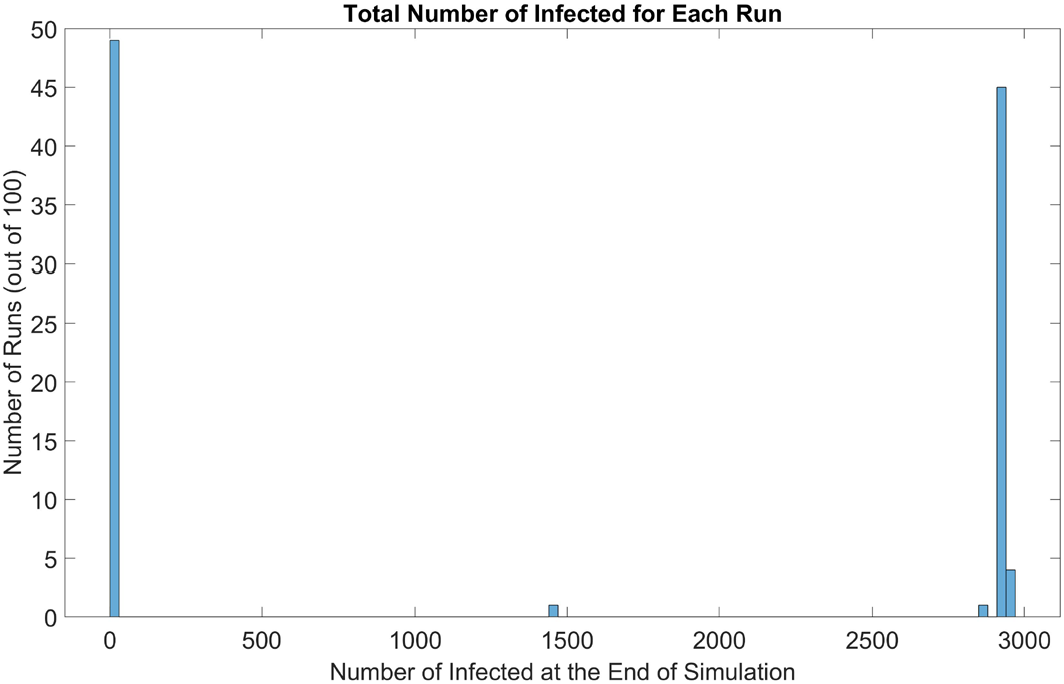
Mode of virus transmission in dormitory setting (baseline, 100 runs). Out of the six modes we modeled, canteens and the hallways of each floor appear to be the safest. Canteens are large enough for students to practice ample social distancing. And hallways see much less traffic than ground floors.

### 3.2 Surgical Mask

In figure 6a, when we increase the percentage of students wearing masks, we observed that the speed of new infections begins to slow down, effectively flattening the curve. In addition, the probability of outbreak also decreases accordingly. When mask usage reaches 75%, the curve starts to linearize; at 100%, it has almost become a horizontal line, greatly slowing down the transmission of COVID-19.

**Figure 6.**
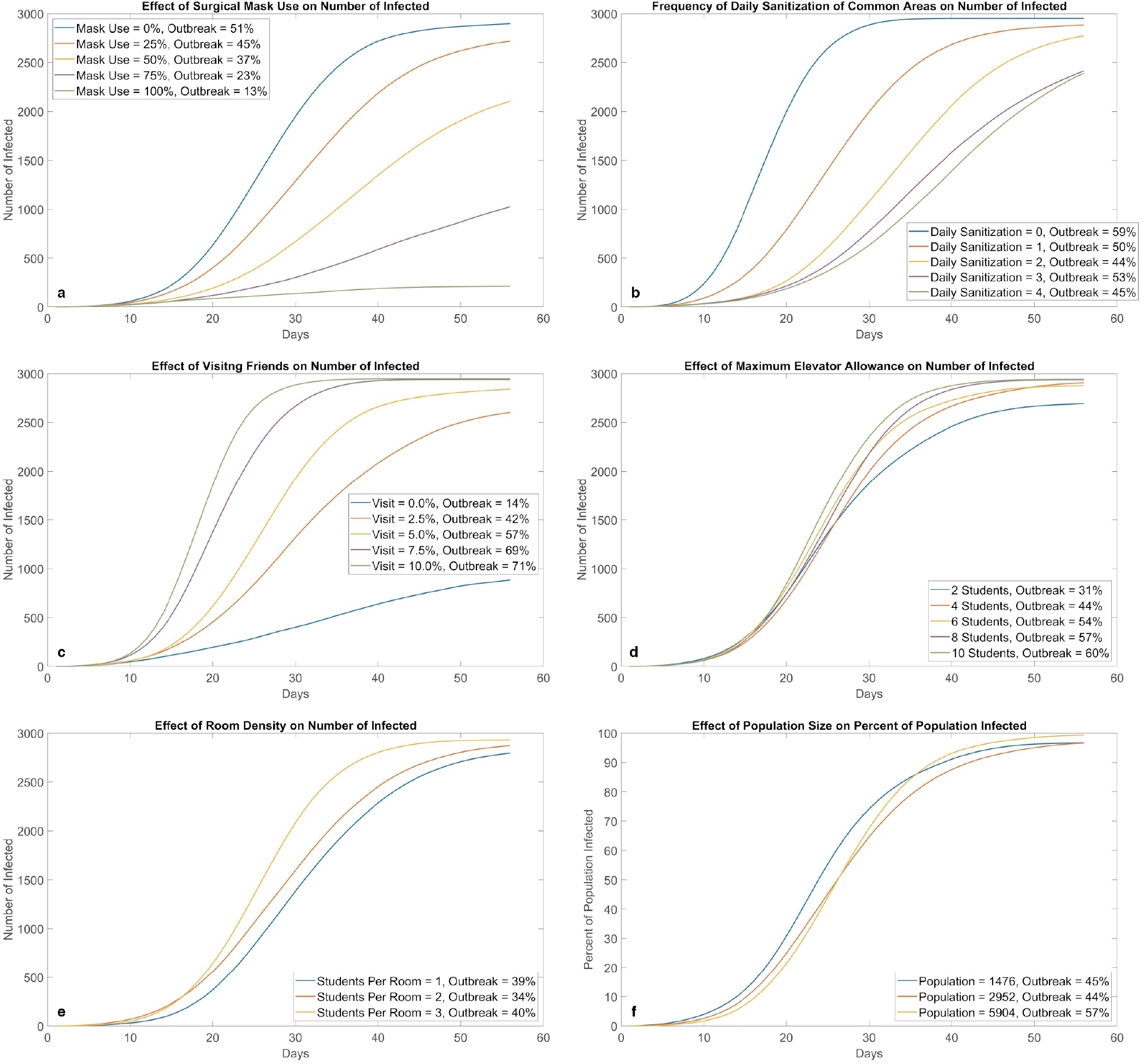
Effects of different NPI strategies (100 runs for each parameter value). Mask adoption (a) and not visiting friends (c) appear to be the most effective at preventing virus transmission on campus; they directly reduce the risk of face-to-face transmission. Increasing cleaning frequency (b) is also effective, however, its potency seems to decrease with each additional cleaning. Density in elevator (d) and density in rooms (e) do not have a large impact flattening the curve. A possible explanation is that these NPIs attempt to reduce the amount of people in the same area during certain time periods, however, the total amount of susceptible students interacting in the system remains the same. A similar trend can be observed for total population (f), where the shape of the curve (with percentage on the y-axis) is relatively insensitive to population changes. This property also holds for deterministic ODE SEIR models.

### 3.3 Sanitization

Figure 6b shows that regularly cleaning all public areas can prevent the spread of COVID-19 with great effectiveness. As a result, sanitizations should be thoroughly conducted as often as possible, as manpower and equipment allow, although the effectiveness of each additional cleaning seems be less and less. In real life, the effectiveness would be lower, since disinfectants do not have a 100% potency like they do in our programming.

### 3.4 Visit

Results in figure 6c show that the more students visit each other, the fastert the infection spreads. Risks are doubled for students who visit others in different building as they need to walk on twice the number of hallways and take twice the elevator rides. When visiting is banned at 0%, we see a drastic flattening of the curve. Students to keep visits to a minimum to protect themselves, their friends, and their roommates. Our results are in accordance with those obtained by Bouchnita and Jebrane^41^ when population movement was restricted by different levels.

### 3.5 Elevator Allowance

Relaxing or tightening elevator allowance at the cost of possible safety or inconvenience, respectively, has very little effect on the shape of the curves (figure 6d). On possible explanation is that the elevator environment can be contaminated; regardless of how many people use elevators at the same time, the total number of people who use elevators are constant, therefore, the number of students at risk of being infected in an elevator remains the same despite restrictions for simultaneous travel.

### 3.6 Density

Even though roommate is the #1 cause of infections in a dormitory, reducing the number of roommates or allocating students into all single rooms did not slow down virus transmission by much as evident in figure 6e. Even without roommates, students can still be infected in other ways.

### 3.7 Population

The curves in figure 6f remain very similar when we half or double the population. In other words, the speed of new infections is directly proportional to the number of students on campus. Further testing of other independent variables at different population levels results in very similar curves and outbreak probabilities as those portrayed in earlier figures. Thus, we conclude that the percentage of population infected overtime is relatively insensitive to changes in population, with or without different interventions. The establishment of this linear relationship means our model can be applied to other populations to predict the spread of COVID-19 with relatively high accuracy.

### 3.8 Optimal Strategy

By combining different NPIs, the risk of COVID-19 transmission in residential students can be drastically reduced. Table 4 summarizes each strategy’s effect on flattening the curve, effect on outbreak probability, and ease to enforce. We also propose an optimal value for each NPI aimed for the safest reopening of campus with 100% student capacity. The performance of this optimal strategy (1000 runs) is compared with that of baseline in figure 7.

**Table 4.** Evaluation on the effectiveness of each NPI (* denotes unchanged from baseline).

|  | NPI Strategy | Variable affected | Optimal Value | Effect on Flattening the Curve | Effect on Outbreak Probability | Ease to Enforce |
| --- | --- | --- | --- | --- | --- | --- |
| 1. | Adopt surgical masks | percent of students who use masks | [100%] | very big<br>●●●●● | very big<br>●●●●● | easy<br>●●●●○ |
| 2. | Sanitize common areas more frequently | frequency of daily sanitizations | [4] | big<br>●●●●○ | very small<br>●○○○○ | medium<br>●●●○○ |
| 3. | Limit unnecessary movements | percent of students who visit friends | [0%] | very big<br>●●●●● | very big<br>●●●●● | medium<br>●●●○○ |
| 4. | Limit max. number of students in an elevator | elevator allowance | [4] | small<br>●●○○○ | big<br>●●●●○ | medium<br>●●●○○ |
| 5. | Limit number of students in each room | density | [3] | medium<br>●●●○○ | very small<br>●○○○○ | very difficult<br>●○○○○ |
| 6. | Limit total number of students on campus | population | [100%] | very small<br>●○○○○ | small<br>●●○○○ | very difficult<br>●○○○○ |

**Figure 7.**
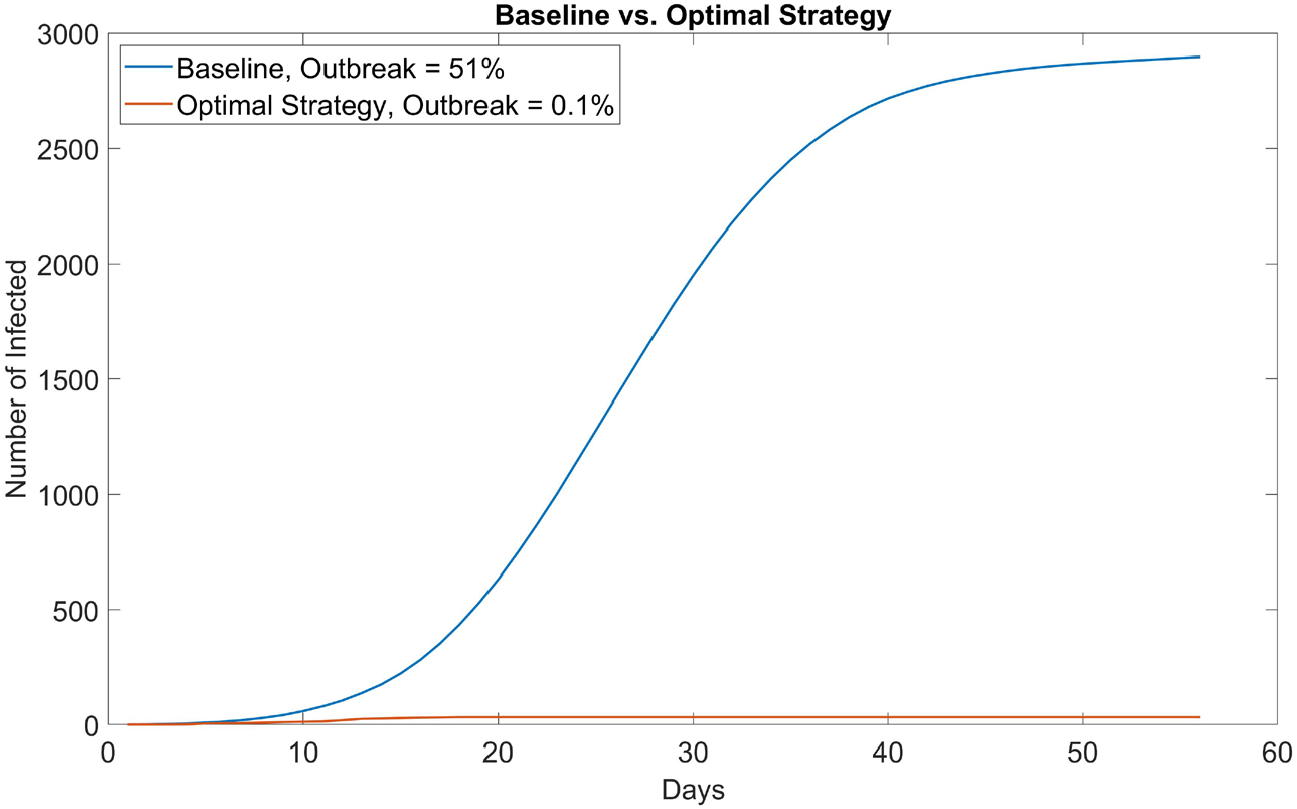
Comparison between baseline no NPI (mask adaptation = 0%, sanitization = 1 time a day, student visiting friends = 5%, max. elevator allowance = 6) and optimal strategy (mask adaptation = 100%, sanitization = 4 times a day, student visiting friends = 0%, max. elevator allowance = 4). In 1000 simulation runs, only one run resulted in outbreak (> 30 infected students); even then, the outbreak size (32 infected) was only slightly above the threshold and eventually contained. School can most likely start with no cases if students properly adopt NPIs and self-quarantine for 14 days before instruction resumes.

## 4. Discussions

With current assumptions, simulation results suggest that universities in low-risk countries can fully reopen even without testing, based on three important premises: 1) all students arrive from low-risk areas where community transmission is minimal, thus minimizing the introduction risk; 2) all students self-quarantine for 14 days on arrival; and 3) all students utilize proper NPIs. Any one singular intervention on its own cannot fully prevent the spread of SARS-CoV-2, therefore, multiple interventions need to be performed to an almost perfect extent, in order to contain this disease. The importance of using a combination of interventions is reiterated by Ghaffarzadegan et al^42^.

Even though NPIs are effective, when cases are detected on campus, it is imperative that the administration take swift actions, such as isolating infected/potentially exposed students, performing contact tracing, and mass testing students in order to halt the initial infection. This aspect is not portrayed in our simulation where we simply let the virus run its course in order to investigate its mode of transmission. The transmissibility *τ* is currently modeled as a constant value of 0.82%. Future work could improve *τ* as a function of time and location to reflect viral discharge and its potency decaying with time^30^ and represent possibly different levels of transmissibility in different environments.

Another key input is the number of initially infected student, denoted as *N*_*1*_. It is dependent on the country and region modeled and cannot be controlled; we can only estimate how many students will arrive infected. Statistically, in a low risk country (our model) where the incidence rate is less than 5 per 100,000 population^13-14^, *N*_*1*_ would most likely be zero. That is, all students will mostly likely arrive healthy, undergo 14 days of self-quarantine, and start in-person instruction with zero infections on campus. However, we included one initially infected student (approx. 0.034% positivity rate) to model the worst case scenario. If other countries are modeled, we would need to update and most like increase *N*_*1*_. For instance, in a university in Indiana, U.S., 33 out of 11,836 (0.28%) students were COVID-19 positive on arrival. Results with *N*_*1*_ = 1 indicate that universities can safely open. Additional research in the future could increase *N*_*1*_ and observe at which value does reopening without testing cease to be a practical and safe option.

To conclude, our model examines a type of university reopening strategy in low risk countries and the transmission of SARA-CoV-2 between residential students on campus. These two areas have not been explored much in previous literatures; however, successful implementation of this strategy can not only safely reopen campus but also potentially save millions of dollars in testing costs. We also selected six different NPIs commonly adopted in school settings and tested how they can assist in the safe reopening of universities. Our results, although based on model assumptions and simplifications, provide useful information for policy makers and campus administrators in countries and areas still struggling with reopening universities during COVID-19.

As with any model, there are certain limitations in our work. Students are programmed to be 100% compliant to protocol; however, this is highly unlikely in real life. They might refuse or forget to wear masks and attend unsanctioned gatherings with friends^8^. These actions will all make them more vulnerable to infection and lead to possible outbreak. In addition, we only modeled residential students and assumed no infection from campus staff, faculty, or outside sources. When 14 days of self-quarantine ends, students are free to go outside. Even though community infection in the city is almost non-existent, it would be interesting to model how the virus propagates through classrooms, athletic events, and other activities when one student is infected from outside or if a certain staff or faculty is infected. Lastly, we did not account for asymptomatic carriers, whose prevalence and infectiousness remain unclear. Based on findings from a number of literatures, the U.S. CDC estimates that 10%-70% of patients could be asymptomatic, and their infectiousness can be anywhere between 25%-100% relative to symptomatic patients^43^. When more epidemiological data are available, future studies could test different prevalence and infectiousness values of asymptomatic patients and examine how they impact the viral transmission in residential students and/or other community settings.

## Data Availability

The NetLogo model is available at: https://www.comses.net/codebases/a5e34ca9-147d-4f9d-aaf7-849a20176946/releases/1.0.0/

## Author Contributions

Chan proposed the initial research idea. Xi and Chan carried out background research to the problem and searched for papers and news reports. Xi created the agent-based model, gathered data, and formulated the paper. Chan provided critical feedback and suggestion.

